# More youthful brain activations observed in older adults with better cognition

**DOI:** 10.1101/2023.07.29.23293355

**Authors:** Yunglin Gazes, Georgette Argiris, Caleb R. Haynes, Yaakov Stern, Christian G. Habeck

## Abstract

We investigated factors that influence the maintenance of youthful functional activation for a working memory task in healthy older adults. While task-related functional activation has been found to increase in extent and intensity in aging, there have been few investigations into the factors that enable more youthful-appearing activation patterns. Similarity to young-like templates underlying more successful cognitive aging is an attractive idea and has been operationalized in a variety of different data domains in neuroscience. We applied an operationalization of such template similarity in a well-understood verbal working-memory task examined in 135 older adults (age=64.9 ± 3.17 years) using 44 younger adults (age=26.0 ± 2.9 years) as the reference group. Topographic similarity to the young-reference pattern was computed as spatial correlation across voxels between a young- and an older-group pattern. Similarity to the young-reference pattern was evaluated via two approaches: group-wise and at the individual level. In group-wise similarity, moderating factors were dichotomized, splitting older adults into two group. Examining the similarity between these group patterns and the young-reference pattern, we observed more youthful-appearing activation patterns for females than males, participants with thicker than thinner cortex, less white matter hyperintensity than more, and better performance in vocabulary, processing speed, memory, and reasoning. No difference in similarity was observed for high vs. low education groups. Individual-level similarity was quantified between the young-reference pattern template and each older adult’s pattern, for which linear regression showed that the degree of similarity to the young pattern was associated with more accurate performance on the working-memory task, even after covarying out age, sex, education, and three brain structural measures. Overall, using a novel approach to quantify topographic similarity between younger and older adults, we provided evidence that older adults with better overall cognition recruit more youthful-appearing activation patterns during working-memory rehearsal, and we discussed its relevance to resilience mechanisms in aging.

## Introduction

Resilience to brain aging is influenced by a variety of factors.^1^ It is still unclear how factors such as sex, structural brain health, and cognitive factors differentially contribute to the maintenance of brain processes in typically aging brains. Functional activation during the performance of a cognitive task, measured with BOLD fMRI, enables examination of the degree and extent of brain regions involved in task processing. Previous studies indicate that older adults recruit more extensive brain regions and with greater intensity than younger adults ^2^. However, the degree to which older adults maintain youthful patterns and the factors that moderate the degree of this differential preservation have not been investigated. Understanding the factors that are associated with greater similarity of older adults’ functional activation to that in younger adults can further our understanding of variability across adults in the cognitive aging process.

Some studies have compared high-vs. low-performing older adults in terms of their similarity to young adults. Duverne el al. ^3^ reported greater voxel-wise differences between young and low-performing older adults than for high-performing adults in functional activation associated with a memory task. In a more direct assessment of young-similarity, Duzel et al. ^4^ quantified the degree of dissimilarity to a young activation pattern for each older adult, using a specially devised scoring scheme called “FADE” score. A newer version of the FADE score, reported in Soch et al.^5^, includes both positive and negative activations and quantifies the degree of similarity rather than dissimilarity. In both versions, greater similarity to the young pattern was associated with better performance in a memory task in older adults.

The primary goal of the current study was to extend this work to test moderating effects on the degree of topographic similarity between older and younger adults’ functional activation patterns, which we will refer to as “young-similarity” for simplicity. Potential moderators included sex, cognitive, and brain structural factors. Rather than using a complex scoring system to operationalize topographic dissimilarity, similarity in functional activation patterns was assessed using a simpler technique of spatial correlations across voxels, which encompassed both positive and negative activations. We examined task-activation fMRI data obtained during a Letter Sternberg verbal working memory paradigm with three load levels (1, 3, and 6 letters)^6^. Two approaches were adapted: group- and individual-level similarity. In group-level similarity, older participants were dichotomized into high- and low-value groups for each potential moderating factor (sex, education, brain structural measures, four cognitive abilities from neuropsychological tests, and performance on the memory task). We then compared young-similarity in these dichotomized groups and generated similarity distributions for group-level activation patterns from resampled data. We hypothesized that greater young-similarity would be observed in the older adult group with more intact brain structural measures and better cognitive-performance measures. In addition to contrasting young-similarity in dichotomized-group-level patterns for older adults, we also quantified individual-level young-similarity in all older adults and examined its association with memory task performance to test the hypothesis that the degree of similarity to the young pattern would correlate with performance on the working memory task after controlling for age, sex, and structural brain measures.

## Results

### Participant characteristics

Analyses included data from 179 strongly right-handed, native English-speaking healthy adults. Sample characteristics and statistics of group difference tests between the younger and older age groups are shown in Table 1. Overall, the older adults showed thinner cortices and greater white matter hyperintensity volume, in line with prior expectations. There were no differences in education or sex distribution between the two age groups.

**Table 1.** Sample characteristics in 179 participants for the current study.

Age associations with brain structural measures
|  |  |  |  |
| --- | --- | --- | --- |
|  | **Younger**<br>**(n=44)** | **Older**<br>**(n=135)** | **Younger = Older?** |
| **Age (M ± STD in years)** | 26.0 ± 2.9 | 64.9 ± 3.2 | P(T-test)<0.0001 |
| **Education (M ± STD in years)** | 15.7 ± 1.9 | 16.1 ± 2.4 | P(T-test)=0.35 |
| **Cortical thickness (M ± STD in mm)** | 2.64 ± 0.11 | 2.50 ± 0.11 | P(T-test)<0.0001 |
| **Sex** | 31 F, 13 M | 75 F, 60 M | P(Fisher exact)=0.11 |
| **WMH (M ± STD in mm<sup>3</sup>)** | 1,285 ± 539 | 2,984 ± 4,162 | P(permutation)=0.0086 |

### Age associations with brain structural measures

Among the older adults, all three of the brain measures, cortical thickness, white matter hyperintensity volume, and white matter mean diffusivity, showed significant correlations with age. Cortical thickness was negatively associated with age: r = -.309, p < .001. White matter hyperintensity volume was positively associated with age: r = .268, p = .002. White matter mean diffusivity, with two subjects missing diffusivity data, also positively correlated with age (higher mean diffusivity = less intact white matter): r = .245, p = .004.

Among the three brain measures, after accounting for age, partial correlation showed that cortical thickness was negatively associated with white matter hyperintensity volume (r = -.176, p = .042) and with mean diffusivity (r = -.204, p = .018), and white matter hyperintensity volume was positively correlated with mean diffusivity (r = .252, p = .002).

### OrT-CVA pattern for Letter Sternberg task activation in the young group

After standard preprocessing and obtaining the task-contrasts, the multivariate Ordinal Trend Canonical Variates Analysis (OrT-CVA) technique was used to derive a group activation pattern for the young participants by identifying voxels that show a monotonic load-related expression (non-decreasing activation from 1, 3, to 6 letter loads) in individual participants. An OrT-CVA pattern with monotonic task-activity curves was found for all but 1 individual in the young group, and a permutation test with 1,000 iterations indicated that the association of the pattern with memory load at the participant level was statistically significant (p=0.002). The resulting pattern served as the reference pattern for subsequent topographic similarity analyses.

### Group-wise topographic similarity to younger adults for dichotomized moderating factors

With the young OrT-CVA pattern as reference, topographic similarity analysis was performed by splitting older participants into two groups based on each of the moderating factors (sex, education, cortical thickness, total white-matter hyperintensity volume (WMH), and neuropsychological and task performance) and calculating the voxel-wise correlation between each group’s activation pattern and the young-reference pattern (young-similarity for short). Permutation test was performed where null conditions are created by swapping people between the high- and low-moderator groups, and the test statistic is the difference between the topographic similarity to the young point-estimate pattern. We obtained the proportion of permuted samples in this null condition that were more extreme than the difference statistics calculated for our unpermuted sample. Random-sampling tests were performed for each moderator to quantify the probability that the group with the higher moderator value (e.g., more white matter hyperintensity) has a functional pattern more like the young pattern. The further the probability value is from 50%, the greater the likelihood of a difference in young-similarity between the two dichotomized groups.

Among the moderating factors examined, only total cognition and episodic memory from neuropsychological tests showed significantly higher degree of young-similarity in functional activation for the high-value than the low-value group. Figure 1 shows the distribution of permuted samples with the vertical line indicating the difference in R for the unpermuted sample for total cognition and for episodic memory. Greater young-similarity was found for the group with better total cognition and better episodic memory than the corresponding lower-performance groups.

**Figure 1.**
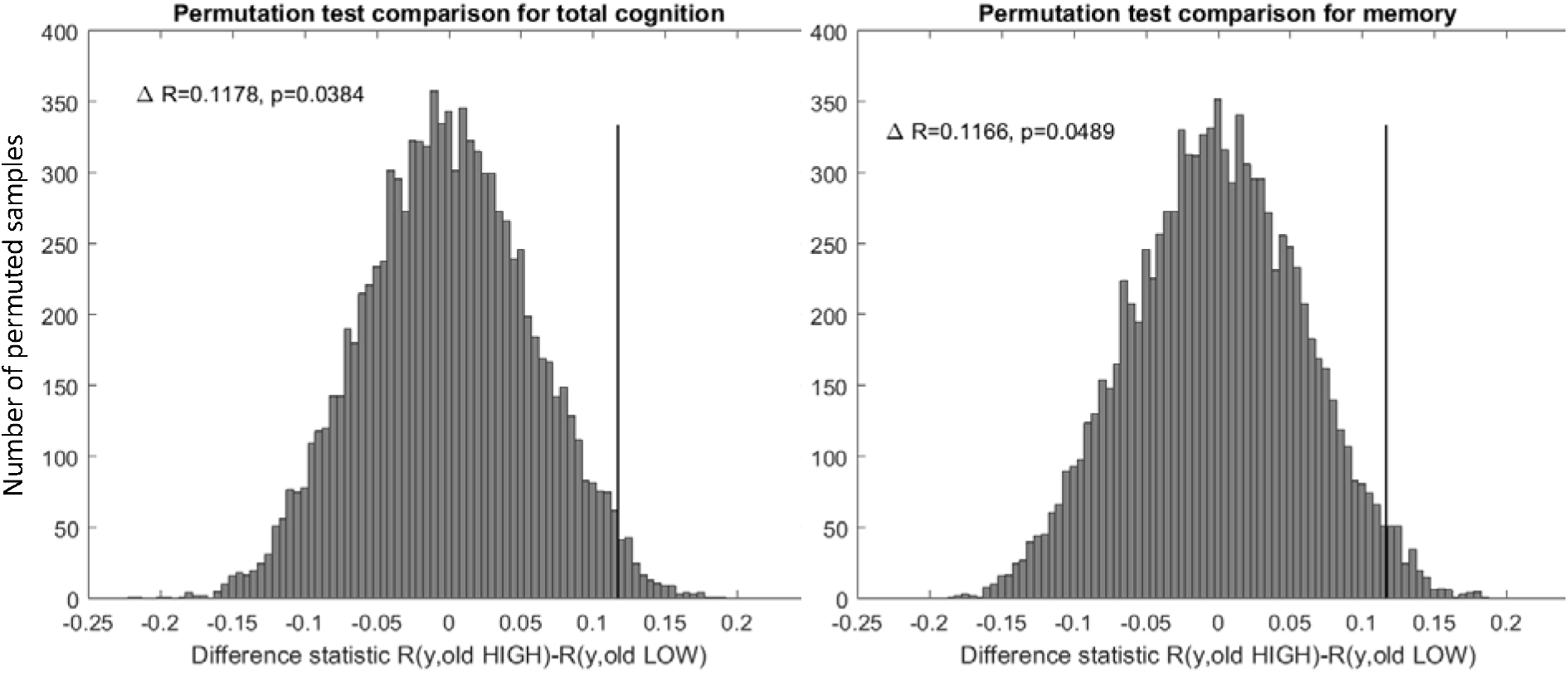
Histograms of permuted samples between young and older adults’ patterns for total cognition and episodic memory. Vertical line indicates the unpermuted sample’s difference between R. The probability, computed from 100,000 random samples, of the higher-moderator-group’s pattern being more similar to the young pattern than the lower-moderator-group is shown above each histogram.

### Associations between individual-level topographic similarity and cognitive performance

In addition to group-wise similarity, topographic similarity to the point estimate of the young reference pattern was also computed for each older individual’s mean task activation map, resulting in a scalar score per individual which was then examined in regression analysis. With age, sex, education, and the three brain measures as covariates, linear regression found a significant association between the topographic similarity as a continuous measure and accuracy on the Letter Sternberg task, demonstrating that more youthful functional activation is associated with better task performance (see Table 2).

**Table 2.**
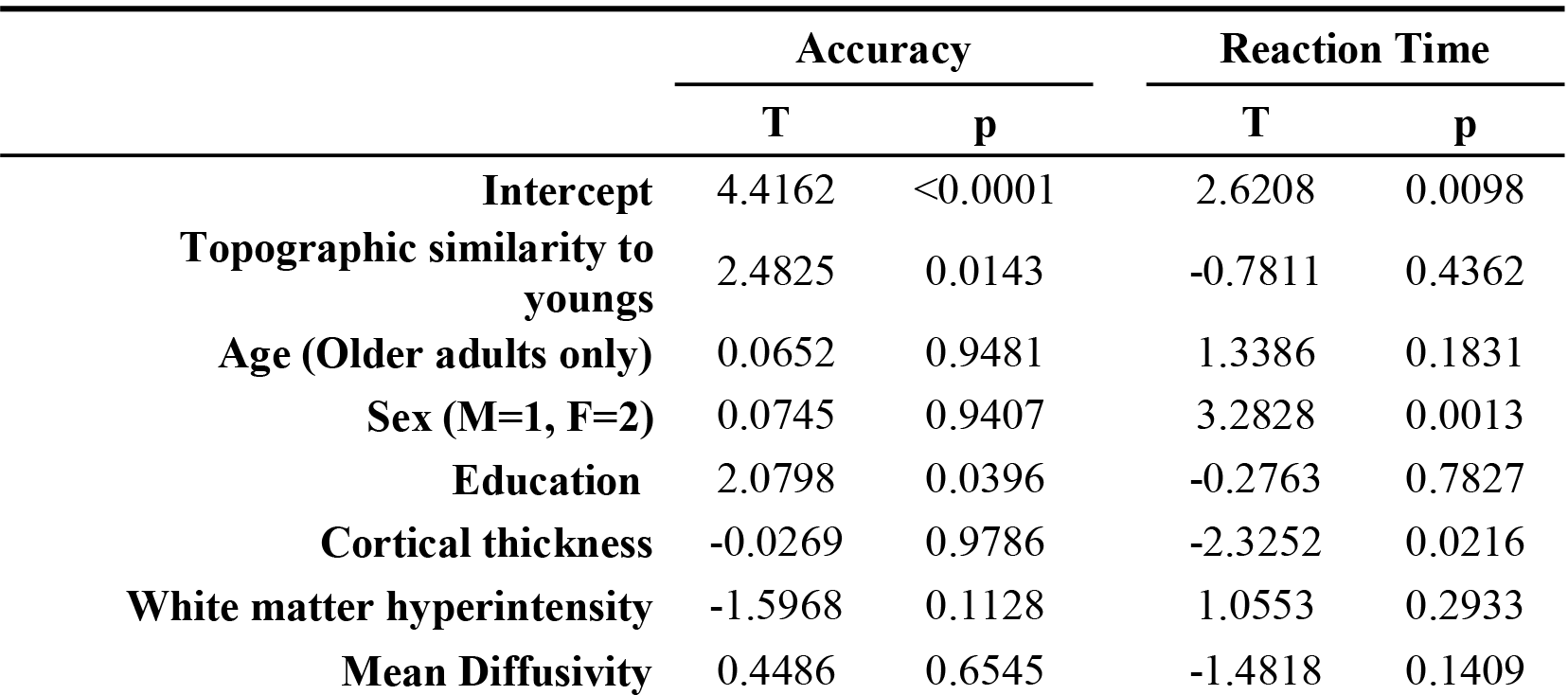
Statistics for regression with accuracy and reaction time as outcomes and topographic similarity, along with other covariates, as regressors.

## Discussion

The current study demonstrated that youthful-appearing activation patterns in older adults are associated with better performance on the fMRI task and for neuropsychological tests. Using a bivariate correlation across voxels for activation patterns on Letter Sternberg verbal working memory task between the young-reference group and each of the two groups for the dichotomized factors, we observed that older adults had greater topographic similarity to the young pattern for better than worse performance on total cognition and episodic memory, which were based on neuropsychological tests. Linear regression also provided evidence of an association between individual-level topographic young-similarity and accuracy on the Letter Sternberg task.

### Template similarity

Using template similarity, our results both replicated and extended the findings of Duzel et al. ^4^. Their measure differed operationally from our simpler measure but can be seen as related. Duzel et al. used visual images as memory stimuli whereas our study used sets of letters. Even though the calculations of topographic similarity and the type of task stimuli were different between Duzel et al.’s and our study, the finding that a more youthful activation pattern is associated with better performance on a memory task was replicated. Extending their findings, our study showed that several factors moderated the degree of young-similarity. While Duzel et al. did not find an association between FADE score and other cognitive scores, our study found that total cognition and episodic memory moderated the topographic similarity of young and older adults’ activation patterns. Furthermore, young-similarity (as an individual-level measure) was associated with task accuracy even after accounting for potential confounders. While this independent association was only correlational, it is consistent with deployment of similar brain regions to the young pattern having beneficial effects for task performance.

Template similarity is conceptually appealing and commonly utilized in cognitive neuroscience. It can be employed in any multivariable data domain, whether it is neuroimaging voxels, neuropsychometric variables, or OMICs data. Despite many different implementation schemes, simple Pearson correlation or covariance computations are most often used and with obvious relevance for multivariate approaches: factor or pattern scores usually encode the similarity of an individual’s multivariate profile with a group-invariant template, and the computation of the factor often involves the covariation of pattern loadings with individual variables in a ‘dot product’ computation. For diagnostic or prognostic applications interested in reducing the multifaceted information from a high-dimensional data set to one scalar measure, similarity measures are an obvious choice, and are usually further transformed into a diagnostic score. However, as in our study, spatial similarity of group-level activation patterns is also of interest, since it captures the similarity of neural processing between participant groups. To bolster the inferential robustness, we applied this similarity measure and generated bootstrap resampling distributions for the group-level patterns in the younger group and the two older groups, beyond a single assessment for point estimates. This enabled assessment of moderating factors on older adults’ degree of young-similarity at the group level.

### Resilience mechanisms against aging

Two mechanisms have been proposed for resilience against detrimental effects of aging: brain maintenance and cognitive reserve^1^. According to the framework established by the Reserve and Resilience Collaboratory (reserveandresilience.com/framework), brain maintenance refers to the “absence of changes in neural resources or neuropathologic change.” Our results demonstrated that individuals with thicker cortices and less white matter hyperintensity show greater young-similarity, which is consistent with the brain maintenance mechanism, even though it is not based on longitudinal data.

For cognitive reserve, the framework specifies that there are neural processes that help cope with age-related brain changes to maintain cognitive performance. Proxies such as estimated IQ (from vocabulary subtest) and lifestyle factors provide an indirect approximation of these processes that are hypothesized to be associated with better development of these cognitive reserve processes. Dichotomization of vocabulary ability in our study showed greater young-similarity for the better performers as hypothesized, which is consistent with the cognitive reserve theory in that older adults with better vocabulary ability also show more youthful activation pattern for a memory task. Since brain structural measures were regressed out, we minimized the influence of brain structural changes, and thus strengthened support for cognitive reserve mechanisms.^1,7^

Maintaining similarity to young participants’ functional processing patterns might be one neural implementation of cognitive reserve, in that it accounts for better performance in the presence of age-related brain changes. Compensation would be an orthogonal concept, in that it leads to reduced similarity while maintaining cognitive performance, hinting at alternate networks that are employed to uphold performance. Our findings in this study are not consistent with the compensatory mechanism.

The lack of young-similarity difference between the high and low education groups, a common proxy for cognitive reserve, is consistent with the growing literature that has failed to show effects of education on a variety of structural and functional outcomes in the context of typical aging ^8-10^. In contrast, education has been associated with better tolerance of AD-related changes^11,12^.

This study suggests a blueprint for the exploration of youthful activation patterns and its relevance for the concepts of cognitive reserve and brain maintenance in aging. While the current results are strictly cross-sectional, a longitudinal extension would shed better light at the temporal sequence of events in structural and functional cognitive aging. Better preservation of youthful activation associated with better concurrent and future cognitive functioning, would manifest one implementation of cognitive reserve. Deviation from young templates and a switch to alternative task-processing networks in higher performers, on the other hand, would manifest compensation, an alternative form of cognitive reserve. Most intriguing would be the relationship to brain maintenance: better preservation of youthful activation patterns could be associated, all things being equal, with higher cortical thickness, reduced white-matter hyper-intensities etc. in the future and vice versa. Our contention is that behavioral lifestyle factors associated with successful aging will be shown to be the underlying mediating mechanisms for preservation of both youthful brain structure and function, and in turn: cognition.

### Moderating factors

While several moderators in our dichotomizing analyses were consistent with expectations, in which greater similarity was observed for better cognition and healthier brain structure, a few moderators warrant further discussion.

Females showed greater young-similarity than males, although this finding had no implications for task performance. Females’ accuracy was not significantly different from males’, while mean reaction time was slower (p=0.002), but young-similarity did not mediate this finding and the finding persisted even when partialing for young-similarity. Even when we created young reference patterns stratified by sex (results not shown), older females showed larger similarity in their group-level patterns to the young females than the older males to the young males [P(young-similarity females > males) = 89.4%]. Hormonal differences between males and females may contribute to the sex difference observed. Research has shown the influence of sex hormones on cognitive performance and brain activation ^13^ and females were shown to have better verbal memory than males ^14^. Despite longer RT in females in our sample, their more youthful activation pattern in the older female adults may demonstrate a manifestation of resilience against aging. With better control of possible confounders, it will be interesting to test more rigorously whether or how biological sex influences the youthful appearance of the neural signature of working memory.

Surprisingly, older participants with higher mean diffusivity showed greater young-similarity than the lower mean diffusivity group. Intact white matter is usually associated with lower mean diffusivity, implying that participants with healthier white-matter structure showed less similarity in their activation patterns to young participants, i.e., resorted to different brain regions. Among the older participants, all three of the brain measures were significantly correlated with each other even after accounting for age, such that a thicker cortex was associated with less white matter hyperintensity volume and lower mean diffusivity. The other two brain measures showed the expected pattern in young-similarity with an association between better brain health and higher young-similarity; thus, it is perplexing that mean diffusivity showed the opposite trend. This finding warrants further investigation.

## Conclusion

This study demonstrated the youthful preservation of functional activation pattern in older adults and examined a number of moderating factors that influence the degree of young-similarity. As expected, more youthful activation patterns were found for older adults with thicker cortices, and less white matter hyperintensities. Further, after adjustment for sex, education, and brain structure, better cognitive abilities and task performance also were associated with higher young-similarity. More interestingly, females showed more youthful activation patterns than males. These findings warrant further investigations with longitudinal data.

## Methods

### Participant sample and demographics

Participants were recruited via random-market-mailing and screened for MRI contraindications or hearing or visual impairment that would impede testing. Older adult participants were additionally screened to eliminate those with dementia or mild cognitive. Other exclusion criteria included: myocardial Infarction, congestive heart failure or any other heart disease, brain disorder such as stroke, tumor, infection, epilepsy, multiple sclerosis, degenerative diseases, head injury (loss of consciousness > 5 mins), mental retardation, seizure, Parkinson’s disease, Huntington’s disease, normal pressure hydrocephalus, essential/familial tremor, Down Syndrome, HIV Infection or AIDS diagnosis, learning disability/dyslexia, ADHD or ADD, uncontrolled hypertension, uncontrolled diabetes mellitus, uncontrolled thyroid or other endocrine disease, uncorrectable vision, color blindness, uncorrectable hearing and implant, any medication targeting central nervous system, cancer within last five years, renal insufficiency, untreated neurosyphillis, any alcohol and drug abuse within last 12 month, recent non-skin neoplastic disease or melanoma, active hepatic disease, insulin dependent diabetes, any history of psychosis or ECT, recent (past 5 years) major depressive, bipolar, or anxiety disorder, objective cognitive impairment (dementia rating scale of <130), and subjective functional impairment (BFAS > 1). A complete description of the participants in terms of demographics and cortical thickness can be found in Table 1.

Informed consent was obtained after study information and potential benefits and risks were fully explained to participants. The study protocol was complied with all relevant ethical regulations and was approved by the Internal Review Board of the College of Physicians and Surgeons of Columbia University.

### Neuropsychological Assessment

All participants completed a standardized battery of neuropsychological assessments, and tasks were administered in the following order: Wechsler Adult Intelligence Scale (WAIS-III) ^15^, Letter-Number Sequencing, American National Adult Reading Test (AMNART) ^15^, Selective Reminding Task (SRT) immediate recall ^16^, WAIS-III Matrix Reasoning ^15^, SRT delayed recall and delayed recognition ^16^, WAIS-III Digit Symbol ^15^, Trail-Making Test versions A and B (TMT-A/B) ^17^, Controlled Word Association (C-F-L) and Category Fluency (animals) ^18^, Stroop Color Word Test ^19^, Wechsler Test of Adult Reading (WTAR) ^20^, WAIS-III Vocabulary ^15^, and WAIS-III Block Design ^15^. Based on a prior analysis in our lab assessing the factor structure of these tasks, four domain scores were generated by z-scoring all tests relative to the full baseline sample, and averaging task z-scores within each domain: Episodic Memory (all SRT outcomes), Vocabulary (WAIS Vocabulary, WTAR, AMNART), Processing Speed (WAIS Digit Symbol, Stroop Color, Stroop Color Word, TMT-A), and Fluid Reasoning (WAIS Matrix Reasoning, WAIS Block Design, TMT-B). The primary outcome measures used in the present study domain z-scores for all cognitive domains. A further average of these four scores was performed to yield a score for total cognition (=G).

### MRI Data Acquisition and Processing

A high resolution structural and fMRI BOLD images of the human brain were acquired in an event-related design using a 3.0 T Philips Achieva Magnet with standard quadrature head coil.

### Structural MRI acquisition and processing

T1-weighted MPRAGE scan was acquired to determine cortical thickness, with a TE/TR of 3/6.5 ms and Flip Angle of 8°, in-plane resolution of 256 x 256, field of view of 25.4 × 25.4 cm, and 165–180 slices in axial direction with slice-thickness/gap of 1/0 mm. Each participant’s structural T1 scans were reconstructed using FreeSurfer v5.1 (http://surfer.nmr.mgh.harvard.edu/). This older version was used to maintain consistency across data for the parent study but with visual inspection already performed for every image, the quality of the parcellations were assured. The accuracy of FreeSurfer’s subcortical segmentation and cortical parcellation ^21,22^ has been reported to be comparable to manual labeling. Each participant’s white and gray matter boundaries, as well as gray matter and cerebral-spinal-fluid boundaries, were visually inspected slice by slice, and manual control points were added in the case of any visible discrepancy. Boundary reconstruction was repeated until satisfactory results for every participant were reached. The subcortical structure borders were plotted by TkMedit visualization tools and compared against the actual brain regions. In the case of discrepancy, they were corrected manually. We took the mean cortical thickness measures reported for both hemispheres and averaged them, to arrive at one global measure per participant.

### White matter hyperintensity

Fluid-attenuated inversion recovery (FLAIR) scan was acquired with the following parameters: 11,000 msec repetition time, 2800 msec echo time, 256x189 voxels in-plane resolution, 23.0x17.96 cm field of view, and 30 slices with slice-thickness/gap of 4/0.5 mm and processed through a fully automatic supervised machine learning technique. This method uses a Randomized Decision Trees algorithm called Random Forest for training of the classifier, which has been shown to be superior to the Support Vector Machine algorithm often used for segmenting WMH. The final segmentation is a probability map in [0, 1], which denotes the likelihood that a given voxel is hyperintense, allowing the calculation per subject of a normalized effective WMH volume. Periventricular and deep hyperintensity accumulations are separated using a ventricular template. Processed WMH images were visually checked and corrected if voxels were erroneously identified as WMH.

### White matter microstructural integrity

Two diffusion MRI were acquired in 56 directions using the following parameters: b = 800 s/mm2, TE = 69 ms, TR = 11,032 ms, flip angle = 90 degrees, in-plane resolution 112 x 112 voxels, acquisition time = 12 min 56 s, slice thickness = 2 mm (no gap), and 75 slices. Diffusion data were analyzed with version 3.0.1 of the software, MRtrix3 (www.mrtrix.org), starting with a set of preprocessing steps to improve data robustness: (1) denoising ^23^, (2) Gibbs ring correction^24^, (3) corrections for motion and eddy currents (FSL eddy)^25^ and (4) bias field correction^26^. Diffusion tensor models were estimated for the preprocessed data from which mean diffusivity (MD) was calculated for each participant. To calculate the mean MD for the white matter across the whole brain, each participant’s T1 structural scan was registered to the mean of the non-diffusion weighted images, on which FreeSurfer parcellation was again performed. The white matter mask derived from FreeSurfer was then used to quantify each participant’s mean MD across all white matter in the brain, resulting in one MD value per participant and used in LCSM described below.

### Letter Sternberg working-memory task

#### Task description

Participants studied an array of either 1, 3, or 6 upper-case letters for 3 seconds (=encoding phase), then rehearsed the letters sub-vocally for 7 seconds facing a blank screen (=maintenance phase), to be probed with a lower-case letter to which they responded with a differential button press whether the letter was part of the studied array or not (=retrieval phase). Task consisted of 3 blocks with 30 trials per block and randomly distributed interleave intervals between trials. For the current study we only focused on the maintenance phase.

#### Data acquisition

Functional data for the task were acquired in 3 runs, each of which included collection of 314 functional volumes using a T2*-weighted gradient-echo echo planar image sequence. 36 transverse slices per volume with 3.0 mm thickness and no gap in between were acquired using a field echo echo-planar imaging (FE–EPI) sequence with the following parameters: TR 2000 ms, TE 20 ms, flip angle 72; in-plane acquisition matrix 112 x 112 matrix; which results in a voxel size 2.0 x 2.0 x 3.0 mm.

Task stimuli were back projected onto a screen located at the foot of the MRI bed using an LCD projector. Participants viewed the screen via a mirror system located in the head coil and, if needed, had vision corrected to normal using MR compatible glasses (manufactured by SafeVision, LLC. Webster Groves, MO). Task administration and collection of behavioral data were conducted using PsyScope 5X B53 ^27^. Task onset was electronically synchronized with the MRI acquisition computer.

#### Pre-processing

FMRIB Software Library v5.0 (FSL) and custom-written Python code were used to perform the following pre-processing steps for each participant’s dataset: All functional images were realigned to the first volume, corrected for the order of slice acquisition, smoothed with a 5 mm^3^ non-linear kernel followed by intensity normalization, and high-pass filtered using a Gaussian kernel and cut-off frequency of 0.008 Hz. For spatial normalization, the accompanying T1-weighted high-resolution anatomic image was co-registered to the first functional volume using the mutual information co-registration algorithm implemented in FLIRT. This co-registered high-resolution image was then registered to MNI standardized space using nonlinear coregistration implemented in FNIRT. These obtained transformation parameters were used to transfer the statistical parametric maps of the subject level analysis to standard space.

#### fMRI subject level analysis

The fMRI time-series data was pre-whitened to explicitly correct for intrinsic autocorrelations in the data. The FEAT module ^28^ in FSL was used for first-level analysis. An event-related design was used to model the fMRI data, allowing us to separate timeouts (where no response was made), correct and incorrect trials, task loads (1,3,6) and task phases (stimulus, retention, probe). Incorrect responses and timeouts were modeled together. For all participants, a first level analysis was run on each of their task-based runs with 9 regressors: 3 task loads x 3 task phases. The regressors were generated by convolving FSL’s canonical double gamma HRF with the duration of the respective task phases: stimulus = 3 sec, retention = 7 sec, probe = RT. A second level analysis was run on each participant by combining the first level contrasts for each run. Contrasts for the retention phase of the three memory loads, 1, 3, 6, were used in subsequent analysis.

### Ordinal Trend Canonical Variates Analysis (OrT-CVA)

A well-established multivariate technique, Ordinal Trend Canonical Variates analysis (OrT-CVA)^29^, was used to generate a group-level activation pattern in the young group. OrT-CVA specifically identifies brain patterns that show a monotonic increase in activation with increasing task load. For the Letter Sternberg task, the OrT-CVA pattern showed an increase in pattern expression during the retention period with greater memory load (1, 3, 6 letters) on an individual-subject level.

We can write the multivariate decomposition achieved by OrT-CVA as follows. The derived activation pattern will be written as v, and participant S’s activation map for memory load L can be written as an indexed column vector y_(S,L)_. This activation map can be written as the product of the group-level activation pattern with a subject- and load-dependent factor score w_(S,L)_ and some unaccounted residual ε:

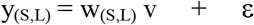

OrT-CVA puts constraints on the factor score w_(S,L)_ and derives a pattern whose factor score shows a positive within-person relationship with memory load for as many participants as possible, with an inferential framework for ascertaining significance through permutation tests, using p < .05 two tailed.

For the estimation of topographic robustness, a bootstrap estimation procedure was conducted which resampled all participants and performed the OrT-CVA point-estimate procedure on the resampled data 100 times. A topographic Z-map was approximated semi-parametrically by computing the bootstrap variability as a standard deviation around the point estimate, and dividing the point estimate by this standard deviation as

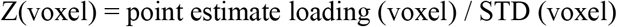

A minimum value of |Z|>2 is demanded for regions to be highlighted with a consistent loading in a visualization plot.

### Topographic similarity to young patterns

#### Approach 1: older adults’ group wise

The focus of the current study was the computation of topographic similarity between the OrT-CVA patterns for younger participants and the corresponding activation patterns for older participants. Since the age distribution is bimodal, ‘older’ and ‘younger’ can be cleanly defined from our age distribution with the younger range of 20 to 30 years which comprised 44 individuals, and the older range of 55 to 70 years which comprised 135 individuals.

For this analysis, we performed OrT-CVA on the younger individuals with a bootstrap resampling procedure that yielded 200 bootstrap patterns and a point-estimate pattern. We then derived similar OrT-CVA patterns in the older subjects, using the following criteria to divide the older subjects into two groups that were split along a median value: education, cortical thickness, total white-matter hyperintensity volume (WMH), and neuropsychological and task performance. For education, cortical thickness, and WMH, we just performed a median split of the older participants. For neuropsychological and task performance, to adjust for the potentially confounding factors, we removed the influence of age, education, sex, cortical thickness, mean diffusivity, and WMH from these measures and then split the residuals along the respective median. Both procedures resulted in an assignment of each older participant into a “high” and “low” group with respect to each variable. For both groups, OrT-CVA was then executed with 200 bootstrap samples.

We then assessed whether the high or low group of the older participants was more like the young group. To do this, we computed all possible 200*200=40,000 pairwise spatial Fisher-Z coefficients between all bootstrap patterns of the young and old-high groups, and again between all bootstrap patterns of the young and old-low groups. The resulting two similarity distributions were then plotted and inspected to see whether difference in the distributions was noticeable.

In addition to computing spatial correlation coefficients for pairs of images across voxels, to visualize the topographic similarity as a voxel-wise map, we also computed voxel-wise “overlap” images. The computation proceeded as follows: all 200*200=40,000 pairings of images between the young group and both high and low-performing older groups were formed, similarly to the computation of the topographic similarity. The loadings of the OrT-maintenance bootstrap patterns (which are always normalized to have a Euclidean norm =1) were multiplied voxel-wise for each pairing, and then summed across all 40,000 pairings (but not across voxels). This procedure resulted in two voxel-wise product-summation images which tallied the similarity to the young reference group on a voxel basis. A high positive voxel value in these images indicates that high loadings with consistent sign at this voxel were multiplied and added up to a large value, whereas a large negative summation value indicates the loadings had a large magnitude but inconsistent signs in the voxel loadings between the young reference group and the older groups. We set an arbitrary threshold to clean the images and decided on a value of 1 for the clearest depiction.

We expected higher-performing older participants to show greater similarity in their maintenance-related activation pattern to the young participants than the lower-performing older participants. Further, we expected older participants with better preserved brains to display higher similarity to the young participants.

### Calculation of probability in young-similarity differences between dichotomized groups

The probability can be approximated that the patterns of young participants and older participations with a higher value of the considered moderator are more similar than the patterns of young participants and older participants with a lower value of the considered moderator. We can randomly sample data points in both similarity distributions 10,000,000 times and compute P (similarity between YOUNG and OLD with high moderator value > between similarity YOUNG and OLD with low moderator value) as a relative frequency.

#### Approach 2: linear regression on older adults’ individual similarity scores

In addition to group-wise similarity, topographic similarity to the point estimate of the young reference pattern was also computed for each older individual’s mean task activation map, resulting in a scalar score per individual. A linear regression analysis on the similarity values with age, sex, education, and the three brain measures as covariates.

## Data Availability

All deidentified neuropsychological and behavioral data, and neuroimaging summary measures, along with analytical scripts used to produce the results presented in the manuscript will be uploaded to Dryad (datadryad.org).

## Data availability

All deidentified neuropsychological and behavioral data, and neuroimaging summary measures, along with analytical scripts used to produce the results presented in the manuscript will be uploaded to Dryad (datadryad.org). Dryad works with Zenodo (zenodo.org) to simplify data sharing by making the data and associated scripts used in a study easily accessible on a public platform. The final http-link will be included in the ultimate manuscript version.

## Code availability

All custom written code used for analysis in the current study will be uploaded to github. The final http-link will be included in the ultimate manuscript version.

